# Evaluating rapid water excitation techniques for 5D whole-heart fat-suppressed free-running cardiac MRI at 1.5T scanner

**DOI:** 10.1101/2025.01.16.25320660

**Authors:** Yasaman Safarkhanlo, Jérôme Yerly, Mariana B. L. Falcão, Adèle LC Mackowiak, Davide Piccini, Matthias Stuber, Bernd Jung, Christoph Gräni, Jessica AM Bastiaansen

## Abstract

**Background:** Balanced steady-state free precession (bSSFP) sequences offer high SNR and excellent tissue contrast for whole-heart MRI. However, inadequate fat suppression can introduce artifacts, particularly with non-Cartesian readouts. This study aimed to compare novel rapid water-excitation (WE) pulses for 3D radial whole-heart free-running MRI at 1.5T, specifically Binomial Off-Resonant Rectangular (BORR), Lipid Insensitive Binomial Off-Resonant RF Excitation (LIBRE), and Lipid Insensitive Binomial Off-Resonant (LIBOR) pulses, alongside a Fast Interrupted Steady-State (FISS) and non-fat suppressed free-running bSSFP sequence.

**Methods:** Three free-running MRI protocols (BORR, LIBRE, LIBOR) were optimized for fat suppression at 1.5T using a phantom. These protocols, along with FISS and non-fat suppressed bSSFP, were tested in phantom and five volunteers, with each acquisition lasting 3min and 41sec.

SNR and CNR_Water-Fat_ were measured in phantom data, while SNR and CNR_Blood-Myocardium_ were assessed in volunteers using static gridded reconstructions. Motion-resolved reconstructions were used for qualitative assessments. SAR values were measured for each sequence, and statistical differences were analyzed using one-way ANOVA (p < 0.05). The pulse with the best combination of high SNR and low SAR was identified for further applications.

**Results:** In phantom studies, LIBOR had the highest CNR_Water-Fat_ (276.8 ± 2.5), followed by LIBRE (268.1 ± 2.6), BORR (249.9 ± 2.2), and FISS (212.7 ± 2.7), though these differences were not statistically significant (p > 0.05). FISS showed the highest SAR (1.41 W.kg^-1^), while LIBOR had the lowest (0.23 W.kg^-1^). In volunteers, BORR had the highest SNR in the ventricular blood pool (17.0 ± 1.5), and LIBRE had the highest CNR_Blood-Fat_ (29.4 ± 9.3). FISS had the highest CNR_Blood-Myocardium_ (29.0 ± 8.9), but the differences were not significant (p > 0.05). LIBOR consistently had the lowest SAR (0.26 W.kg^-1^).

**Conclusion:** This study compared various fat-signal-suppression approaches in 3D contrast-free whole-heart free-running bSSFP MRI at 1.5T. Although the sequences performed similarly in SNR and CNR, LIBOR offered the lowest SAR, making it a promising candidate for future whole-heart MRI applications, particularly where RF energy deposition is a concern.

## INTRODUCTION

Double oblique 2D imaging in cardiac MRI (CMR) typically necessitates skilled technicians for precise planning and requires cardiac triggering and patients to perform multiple breath-holds [1]. Free-running whole-heart MRI greatly simplifies the acquisition process and eliminates the need for breath-holds [2], both of which are highly desirable in clinical settings. Radial trajectories [3] are inherently more robust to cardiac and respiratory motion compared to Cartesian trajectories, which enhances the overall image quality [4]. At 1.5T, the free-breathing contrast-free sequence employs a spiral phyllotaxis trajectory [3], which allows for the retrospective reconstruction of motion-resolved 5D whole-heart sparse MRI images [5]. However, radial trajectory readouts can lead to inconsistent and less uniform k-space coverage, which complicates the application of conventional fat suppression techniques, such as a chemical shift-based method like Dixon. As a result, water-excitation approaches are often the preferred method for fat signal suppression in non-Cartesian trajectories [6,7].

To achieve a high signal-to-noise ratio (SNR) and excellent blood-myocardium tissue contrast without using contrast agents [8], it is ideal to acquire a free-running whole-heart MRI with a balanced steady-state free precession (bSSFP) sequence [9,10]. However, bSSFP imaging produces bright water and fat signals, which can impede the differentiation between pericardial fluid and fat, blurring the visualization of small anatomical structures, such as coronary arteries, and increasing artifacts from surrounding fat. Additionally, bSSFP may introduce fat-water cancellation artifacts, further obscuring vessels and other fat-water interfaces. To overcome these limitations, fat signal suppression techniques can be used, including chemical selective fat suppression [11], short inversion time inversion recovery [12], or specific magnetization preparation modules [13,14]. However, these methods may lead to longer repetition times (TR) [16] or introduce sensitivity to B0 inhomogeneity and partial-volume effects [17]. Nonetheless, inadequate fat signal suppression in bSSFP sequences can lead to India ink artifacts [2,18] and compromise the overall image quality by introducing streaking artifacts [6] for radial trajectories, particularly in cases where subcutaneous chest fat is not effectively suppressed. Due to the fast T1 recovery of fat signal and the k-space center dominated MRI signal weighting, achieving effective fat suppression becomes particularly challenging in advanced whole-heart MRI techniques that employ non-Cartesian readouts [19,20]. In these cases, water excitation (WE) methods are generally favored over fat saturation methods for such readouts, as they selectively excite water without relying on uniform fat suppression [6,21]. However, at lower magnetic field strengths, the resonance frequency difference between water and fat narrows, which increases the WE pulse durations and prolongs the overall acquisition time.

The first use of a conventional fat suppression pulse [11] in the free-running bSSFP approach for whole-heart imaging at 1.5T scanners required an acquisition time of up to 14 minutes to comply with specific absorption rate (SAR) limitations [2]. To address this limitation, Fast Interrupted Steady-State (FISS) sequences were introduced [22–24], which interrupt the bSSFP steady state to create a wide signal stop-band centered around the main fat resonance. However, this technique restricts TR to a narrow range, which is essential for optimal fat suppression. This restricted TR range, while beneficial for fat suppression, limits the flexibility of the sequence and directly impacts the achievable spatial resolution, making it more challenging to produce high-resolution images [24]. For example, at 1.5T, FISS requires a TR range of 2.2 - 3.0ms with 8 readouts per FISS module, which is well-suited for anatomical imaging applications that do not demand high spatial resolution [24,25]. To address the lengthy acquisition times and the extended duration of conventional water excitation pulses, off-resonant water excitation (WE) pulses have been proposed, such as the Binomial Off-Resonant Rectangular (BORR) [26] and the Lipid Insensitive Binomial off-Resonant RF Excitation (LIBRE) [6,27]. These RF pulses have a relatively short pulse duration at the expense of an increased RF power [23,27–29], resulting in a higher RF energy deposition. However, they significantly improve fat signal suppression and have been applied in various imaging applications [6,7,30–36]. Incorporating such pulses in a bSSFP sequence with an intensive SAR can present practical challenges. For instance, in a whole-heart free-running bSSFP acquisition at a 1.5T scanner, the optimal contrast between the blood pool and myocardium was compromised by the SAR limitations when the LIBRE pulse was used [7], highlighting the difficulty of balancing effective fat suppression and image quality with the need to adhere to SAR restrictions, particularly in longer acquisitions.

Among the mentioned off-resonant water excitation pulses, BORR provides a broad fat suppression bandwidth but at the cost of higher SAR, whereas LIBRE offers shorter pulse durations and improved efficiency. However, an RF-power-optimized off-resonant water excitation pulse, called Lipid Insensitive Binomial Off-Resonant (LIBOR) pulse, was proposed for respiratory-self-navigated whole-heart free-running MRI at 3T [29], combining these strengths by optimizing RF power and frequency offsets, resulting in lower SAR and robust fat suppression. This makes this pulse particularly suitable for SAR-intensive acquisitions, such as those using bSSFP sequences. Additionally, the LIBOR pulse offers flexible RF duration, which can be shortened like LIBRE and BORR. Given these characteristics, the potential of LIBOR pulses for the whole-heart free-running bSSFP sequence at a 1.5T scanner is promising, as it simultaneously tackles both SAR minimization and fat signal suppression challenges.

Fat suppression remains a critical challenge for 3D radial bSSFP sequences, particularly in whole-heart imaging at clinical 1.5T scanners. Despite various fat suppression approach developments, they have not yet been optimized or systematically compared in free-running whole-heart bSSFP sequences [29]. Therefore, this study aimed to implement the LIBOR WE pulse for non-contrast free-running whole-heart MRI at 1.5T and perform a comprehensive comparison with water excitation BORR and LIBRE pulses, and the fat-signal suppression technique, FISS. The fat suppression capabilities, SAR deposits, and other relevant metrics were quantified for each method in phantom and healthy volunteers’ experiments. By doing so, this study tried to identify the method that achieves the highest SNR while minimizing SAR, ensuring optimal image quality for clinical applications.

## METHODS

### Implementation of water excitation pulses for the 3D radial free-running bSSFP research sequence

BORR, LIBRE, and LIBOR are the main available off-resonance WE pulses for the 3D radial free-running research sequence. These pulses follow a binomial pattern where the first and second RF subpulses are spaced to selectively excite water while leaving fat mostly unexcited, similar to the approach used in the 1-90°-1 water excitation pulse. Additionally, these pulses incorporate phase modulation on the second RF subpulse, a technique used to adjust the phase of the RF signal to better target specific frequencies, such as water. This modulation reduces the total pulse duration by half compared to the conventional 1-180°-1 water excitation pulse, though it comes at the cost of reduced fat suppression bandwidth (BW) [29]. This shorter pulse duration improves time efficiency at the cost of slice selectivity, meaning the pulse affects a larger volume of tissue, potentially introducing signals from outside the desired area and leading to artifacts in certain imaging scenarios. However, while LIBRE generally allows for shorter pulse durations than BORR due to its optimized design, in this study, all three pulses (BORR, LIBRE, and LIBOR) were standardized to a pulse duration of 1.3 ms. This was done to ensure consistency and fair comparison of their fat suppression performance under identical acquisition conditions.

The BORR pulse incorporates a 180° phase modulation on the second subpulse with an RF excitation frequency modification. Theoretically, it can provide a wide fat suppression band; however, its performance has not yet been tested with a 3D radial free-running bSSFP sequence on 1.5T scanners.

The LIBRE pulse offers a shorter pulse duration, outperforming conventional fat suppression methods and water excitation approaches. This pulse has been used for the 3D radial free-running MRI at 1.5T and 3T scanners [6,7,27].

The LIBOR pulse has an excitation frequency of 270 Hz, half that of the LIBRE pulse, to enhance the pulse’s efficiency in exciting water while lowering the required RF power (**Table 1)** [30]. Similar to the work by Lin et al. [37], LIBOR’s second sub-pulse requires a specific phase offset to achieve optimal fat suppression, determined through Bloch equation simulations and validated in experimental settings at 3T [29]. This study aims to implement the LIBOR pulse available for the 3D radial free-running bSSFP sequences at 1.5T by optimizing its parameters and testing its performance.

**Table 1.**
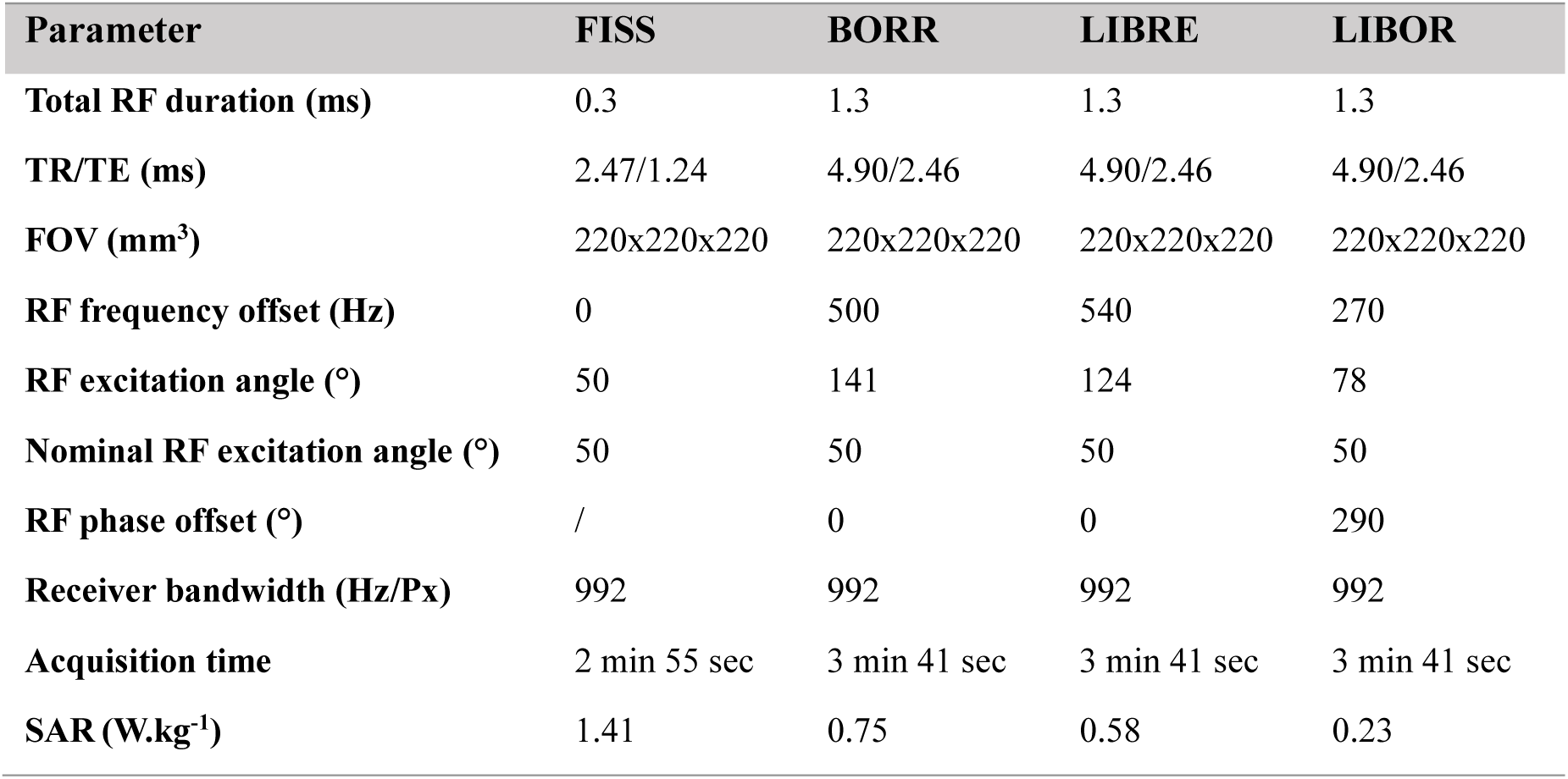
RF excitation properties with their SAR values in phantom experiments. Four different acquisitions were conducted in this study: FISS, BORR, LIBRE, and LIBOR. Each scan was optimized with tuned pulse parameters, including RF frequency offset, excitation angle, and phase offset, to achieve the best fat suppression performance. The nominal RF excitation angle refers to the rotation angle of the water magnetization. The RF excitation angle represents the value displayed and adjusted by the user in the scanner interface.

| Parameter | FISS | BORR | LIBRE | LIBOR |
| --- | --- | --- | --- | --- |
| Total RF duration (ms) | 0.3 | 1.3 | 1.3 | 1.3 |
| TR/TE (ms) | 2.47/1.24 | 4.90/2.46 | 4.90/2.46 | 4.90/2.46 |
| FOV (mm<sup>3</sup>) | 220x220x220 | 220x220x220 | 220x220x220 | 220x220x220 |
| RF frequency offset (Hz) | 0 | 500 | 540 | 270 |
| RF excitation angle (°) | 50 | 141 | 124 | 78 |
| Nominal RF excitation angle (°) | 50 | 50 | 50 | 50 |
| RF phase offset (°) | / | 0 | 0 | 290 |
| Receiver bandwidth (Hz/Px) | 992 | 992 | 992 | 992 |
| Acquisition time | 2 min 55 sec | 3 min 41 sec | 3 min 41 sec | 3 min 41 sec |
| SAR (W.kg<sup>-1</sup>) | 1.41 | 0.75 | 0.58 | 0.23 |

### Water excitation pulse optimization in a fat phantom

MRI experiments were performed using a 3D radial free-running bSSFP MRI research sequence with different off-resonance water excitation methods on a 1.5T clinical MRI scanner (MAGNETOM Sola, Siemens Healthcare, Erlangen, Germany). Each experiment setup was repeated three times over three separate days to ensure consistency. The phantom used for these studies contained vials with different fat percentages, ranging from 0% to 100% of water and peanut oil mixture. The water and peanut oil mixtures were stabilized by immersing the vials in a 3% weight agar solution, which ensured consistent suspension of the components throughout the experiments. Peanut oil was chosen for its resonance frequencies, which closely match those of triglycerides in human adipose tissue, as previously validated [38]. Distilled water was used in the mixtures to minimize impurities and ensure consistency. To further verify the fat content and validate the phantom setup, unlocalized ^1^H MRS measurements were performed in prior work using a high-field MRI scanner, confirming the accuracy of the fat-water compositions used [38].

The study included two independent sets of experiments. First, RF pulse parameters — such as RF excitation frequency, excitation angle, and phase offset—were optimized for each water excitation pulse (BORR, LIBRE, and LIBOR) independently for contrast between water and fat to ensure a fair comparison. In the second experiment, the effectiveness of fat signal suppression for every pulse was compared to each other using the optimized RF parameters from the first experiment. The FISS sequence was also included in this comparison, with its parameters taken from previous studies [24] where they had already been optimized, so no further adjustments were required for this experiment.

To find the optimal values, each RF parameter was systematically changed one at a time within a specific range, based on previous studies using LIBRE at 1.5T [7] and prior work on BORR, LIBRE, and LIBOR at 3T [29]. The initial values for setup were drawn from the mentioned previous literature to ensure consistency with prior work, and each parameter was adjusted independently to ensure accurate optimization for each pulse sequence.

During the optimization of RF excitation frequency offsets, the RF excitation angle was kept constant. Frequency offsets for BORR, with a fixed RF excitation angle of 141 degrees, ranged from 400 to 600 Hz in steps of 20 Hz increments. For LIBRE, with an RF excitation angle of 124 degrees, the same frequency range was used. For LIBOR, with an RF excitation angle of 78 degrees and a phase offset of 285 degrees, frequencies ranged from 250 to 340 Hz in 10 Hz increments.

To optimize the RF excitation angle, the RF excitation offset was held constant. For BORR, with a frequency offset of 600 Hz, RF excitation angles were varied in 10-degree increments from 90 to 180 degrees. For LIBRE, with a frequency offset of 500 Hz, angles ranged from 50 to 180 degrees in steps of 10 degrees. For LIBOR, with a frequency offset of 270 Hz and a phase offset of 285 degrees, RF excitation angles ranged from 50 to 100 degrees in steps of 5 degrees.

Finally, a phase offset sweep from 250 to 340 degrees, with steps of 10 degrees, was conducted for LIBOR pulse with a frequency of 270 Hz and RF excitation angle of 78 degrees to determine the optimal value for achieving the maximum water-fat contrast.

Other key acquisition parameters were kept constant across all scans, with a field of view (FOV) of 220 x 220 x 220 mm^3^, 2.0 mm^3^ isotropic resolution with a matrix size of 112 x 112, 992 Hz/pixel bandwidth (BW), 24 lines per interleaf with 246 interleaves for a total of 5904 radial readouts, 2.46 ms echo time (TE), and TR of 4.9 ms. The acquisition time for each measurement was approximately 30 seconds. Modifications to the scanner console’s user interface enabled the selection between different RF pulses and adjusting the RF excitation frequency, RF excitation angle, and phase offset modulation of the second sub-pulse.

After finding the optimal RF parameters for each pulse, the second experiment for comparing different methods was performed with a higher number of radial readouts (24 x 1877) and the same setup from the RF parameters optimization experiment for the BORR, LIBRE, and LIBOR pulses. These methods all had a constant acquisition time of 3 minutes 41 seconds. These pulses were also compared to a FISS sequence with previously optimized acquisition parameters, summarized in **Table 1**.

### Contrast-free 5D whole-heart free-running MRI in volunteers

Five healthy volunteers (n=5; 4 females; 27.4 ± 7.5 years old; height 168.8 ± 7.8 cm; weight 64.4 ± 10.9 kgs) participated in this study, which provided written and informed consent approved by our local ethical review board.

The volunteer’s protocol was based on the ECG-triggered non-interrupted 3D radial free-running bSSFP sequence, as used in the comparison setup for the phantom experiment. Data were acquired on a 1.5T clinical MRI scanner (MAGNETOM Sola, Siemens Healthcare, Erlangen, Germany) with four fat suppression methods (FISS, BORR, LIBRE, and LIBOR), along with a bSSFP sequence without any fat signal suppression. This comparison followed a similar setup described in a previous study by Masala *et al.* [7], where a 32-channel spine coil and an 18-channel chest coil were used during the free-breathing acquisition. The radial readouts were arranged in interleaves based on a spiral phyllotaxis pattern, with successive interleaves rotated about the z-axis by the golden angle, and the first readout in each interleave oriented in the superior-inferior (SI) direction for subsequent physiological motion extraction and binning into cardiac and respiratory phases.

Acquisition parameters for the sequences were carefully matched to those used in the phantom study for the fat suppression methods comparison (Table 1), ensuring consistency across both setups. In free-running MRI sequences, the acquisition time for each sequence was kept constant for all volunteers, independent of their respiratory pattern and heart rates, with 3 minutes and 41 seconds for BORR, LIBRE, and LIBOR, 2 minutes and 55 seconds for FISS, and 1 minute and 51 seconds for the bSSFP sequence without fat suppression. Although both the acquisition times and repetition times varied between sequences— TR of 2.47 ms for bSSFP and FISS, and 4.9 ms for LIBRE, LIBOR, and BORR—the number of radial lines acquired was kept the same across all sequences, with each acquiring 24×1877 radial lines. This ensured that the data volume and sampling density were consistent despite the differences in TR and total acquisition time. As a result, the comparison between sequences remained valid, with the consistent number of radial lines ensuring comparable data collection.

### 5D whole-heart motion resolved volunteer image reconstruction

Volunteer data were reconstructed using an in-house MATLAB code, following the free-running framework described by Di Sopra *et al.* [39]. Two separate reconstructions were performed: First, a static gridding reconstruction to maintain the noise properties for quantitative analysis, specifically for SNR and contrast-to-noise ratio (CNR) comparisons. This reconstruction used radially sampled k-space data with coil sensitivity combination, implemented using the gpuNUFFT library. Second, a compressed sensing reconstruction was carried out to generate 5D cardiac and respiratory motion-resolved images. This was achieved using the Alternating Direction Method of Multipliers (ADMM) algorithm [39,40].

To enhance the quality of image reconstruction and improve local low-rank (LLR) representations, several regularization techniques are possible, including total variation in the cardiac (TVt), respiratory (TVr), and spatial (TVs) dimensions, as well as local low-rank constraints on cardiac (LRtWeight) and respiratory dimension (LRrWeight). The regularization weights were optimized through a grid search to achieve the best compromise between preserving image detail and minimizing noise. The selected weights were set as follows: TVsWeight = 0, TVtWeight = 0.01, TVrWeight = 0.01, LRtWeight = 0.05, and LRrWeight = 0.05. This combination effectively suppressed artifacts and noise while maintaining both the spatial and temporal resolution of the reconstructed images, which were then used for qualitative assessments of cardiac and respiratory motion.

The motion-resolved reconstruction included 4 respiratory phases and 25 cardiac phases, allowing detailed visualization of both cardiac and respiratory motion throughout the imaging volume.

Reconstruction time for each dataset was approximately 3 hours, executed using MATLAB R2022.b on an Ubuntu 22.04.3 workstation equipped with two 32-core CPUs (AMD Ryzen Threadripper 2990WX, Santa Clara, CA), 128 GB of RAM, and an NVIDIA GeForce RTX 2080 Ti (Nvidia, Santa Clara, CA).

### Data analysis

For the phantom data analysis, DICOM images reconstructed directly at the scanner were used. Analysis followed a structured approach to ensure consistency and reliability across all acquisitions. The SNR was calculated in selected regions of interest (ROIs) on the same fat fraction vial in each acquisition, ensuring consistent relative positioning across all images. Signal values were measured in three selected ROIs from water and fat vials, while standard deviations (SD) were recorded for the background noise. Noise SD was measured by selecting a region in the background of the image, devoid of any object or visible artifacts, to ensure that it accurately represented the baseline noise level. This region was chosen consistently across all acquisitions to maintain reliability. SNR was then calculated as the ratio of the mean signal within the ROI to the SD of the background noise. All measurements were conducted using ImageJ software (National Institutes of Health, Wisconsin University, Bethesda, MD), with uniform brightness and contrast settings applied to maintain consistency across images. For CNR calculations, the difference in SNR between two distinct ROIs was computed.

In the volunteer studies, SNR and CNR calculations were based on 3D static gridding reconstruction to retain original noise properties. SNR was measured using ROIs drawn in the septal myocardium, left ventricular blood pool, chest fat, right lung, and background, in the axial view being visually chosen for ROI placement. The CNRs were calculated by subtracting the blood pool SNR from the chest fat SNR (CNR_Blood-Fat_) and myocardium SNR (CNR_Blood-Myocardium_). For the 5D motion-resolved images, only qualitative assessments were performed, as the compressed sensing (CS) reconstruction altered the noise characteristics, rendering standard SNR and CNR measurements unsuitable. Since these analyses focused on objective, quantitative measurements rather than subjective evaluations, no additional blinding to pulse sequences was deemed necessary.

For both phantom and volunteer studies, Specific Absorption Rate (SAR) values, representing RF energy absorption levels, were retrieved from the DICOM files. These values were compared between different acquisitions to evaluate the relative RF energy absorption levels.

### Statistical analysis

All results are reported as means ± standard deviations. A one-way analysis of variance (ANOVA) was conducted to evaluate differences in SNR and CNR between the pulse sequences, with statistical significance set at p < 0.05. If significant differences were detected, Tukey’s Honestly Significant Difference (HSD) test was applied as a post-hoc analysis to identify pairwise differences while controlling for Type I errors.

SAR values were measured for each sequence and reviewed separately to assess RF energy absorption. The optimal pulse sequence was selected based on a combined assessment of high SNR and low SAR. The sequence demonstrating the best balance between these two factors was identified for further applications.

## RESULTS

### Fat suppression pulse performance in phantom experiments

After performing the experiments three times over three separate days, the RF parameter settings for each pulse consistently converged to the following values to achieve the maximum CNR between water and fat. For LIBOR, the optimal parameters were an RF excitation frequency of 270 Hz, a 70-degree RF excitation angle, and a 290-degree phase offset (**Figure 1**). For BORR, the highest CNR was obtained with a 500 Hz excitation frequency and an RF excitation angle of 140 degrees, while for the LIBRE pulse, the optimal settings were a frequency of 540 Hz and a 120-degree RF excitation angle. The performance of each pulse acquisition in the phantom experiments was visually comparable, with similar brightness levels observed across all acquisitions in vials containing different fat percentages for each pulse sequence (**Figure 2**).

**Figure 1.**
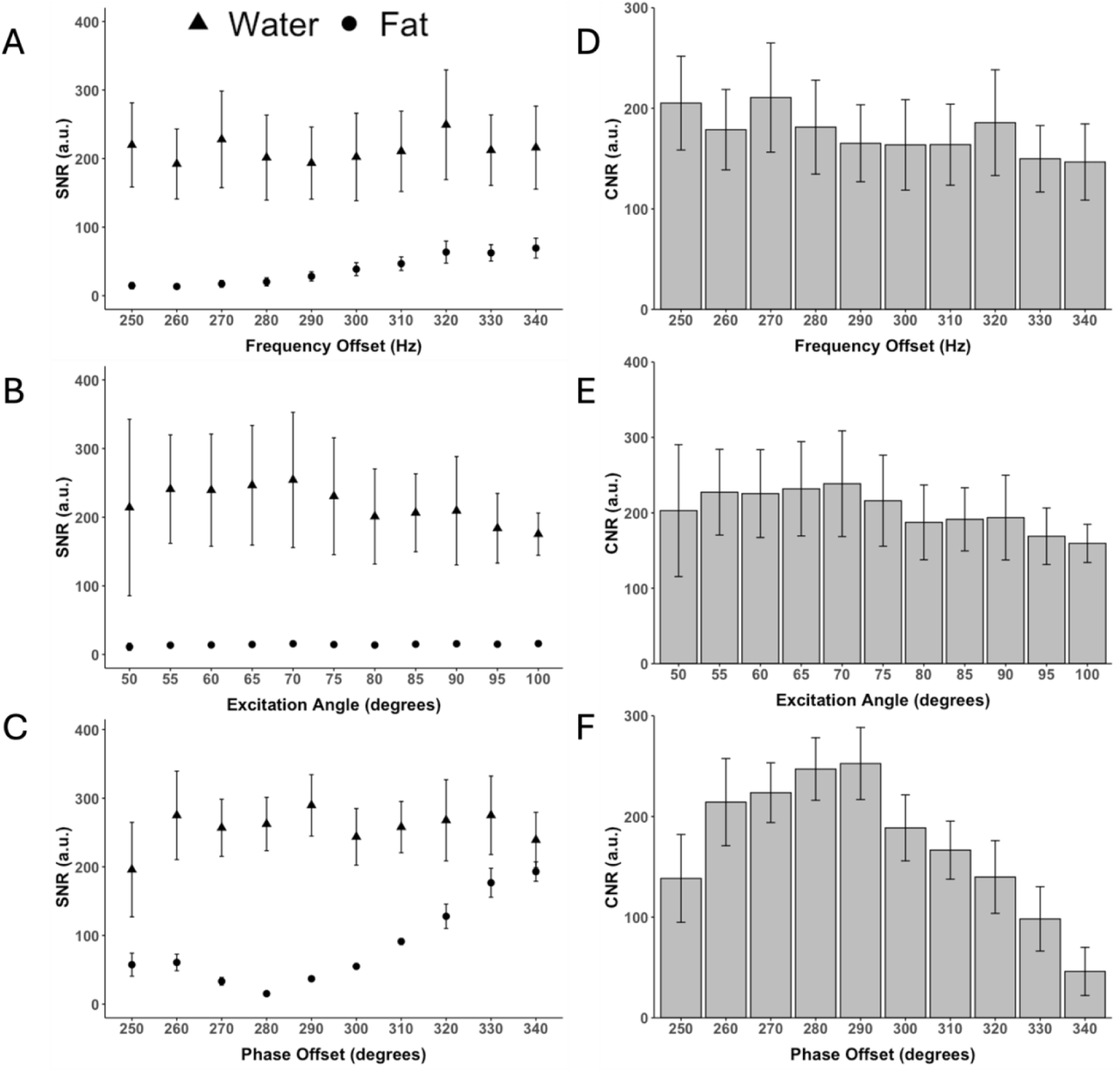
Fat and water signal to noise (SNR) and contrast to noise (CNR) ratios for LIBOR pulse across varying RF parameters optimization in a fat phantom. The SNR for (A) RF excitation frequency offsets (fixed phase offset 285° and a 78° RF excitation angle), (B) RF excitation angles (fixed offset phase of 285° and frequency of 270Hz), and (C) phase offsets (fixed frequency offset of 270Hz and a 78° RF excitation angle). Water-fat CNR for different (D) frequency offsets, (E) RF excitation angles, and (F) phase offsets. Dots denote fat, and triangles represent water.

**Figure 2.**
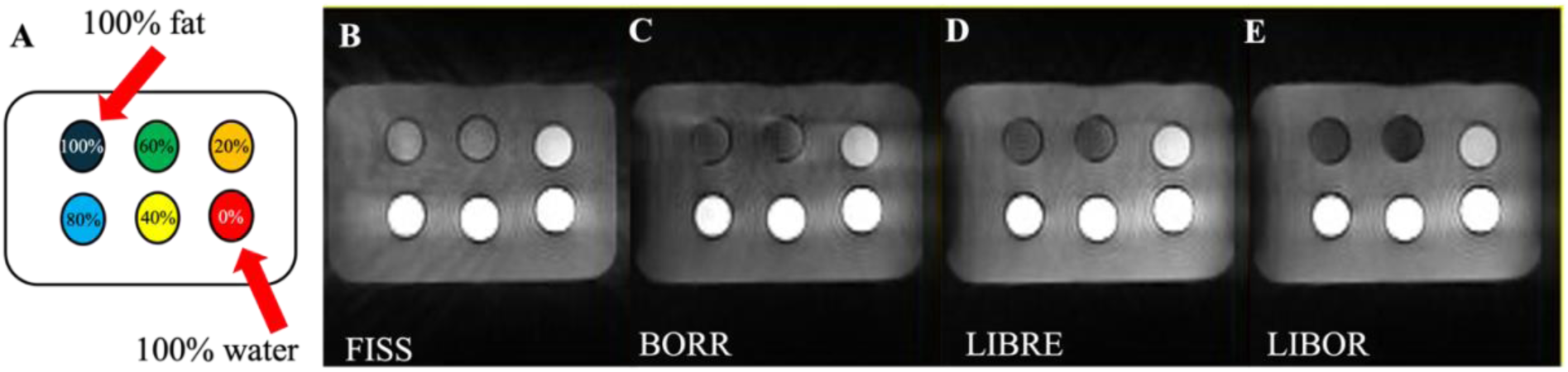
Comparative water-fat contrast assessment using various fat suppression methods, FISS, BORR, LIBRE, and LIBOR, in a phantom experiment. (A) Phantom configuration featuring a 100% fat vial in the top left and 100% water in the bottom right corner, highlighted by the red arrow. (B) FISS pulse with an RF excitation angle of 50 degrees. (C) BORR pulse with a frequency of 500 Hz and an RF excitation angle of 141 degrees. (D) LIBRE pulse with a frequency of 540 Hz and an RF excitation angle of 124 degrees. (E) LIBOR pulse with a frequency offset of 270 Hz, an RF excitation angle of 78 degrees, and a phase offset of 290 degrees. Contrast window and level settings are the same in all images.

Comparing CNR between water and fat for different fat suppression pulses, LIBOR (276.8 ± 2.5) exhibited the highest value, while FISS (212.7 ± 2.7) had the lowest. The CNR values for BORR and LIBRE were 249.9 ± 2.2 and 268.1 ± 2.6, respectively. (**Figure 3**).

**Figure 3.**
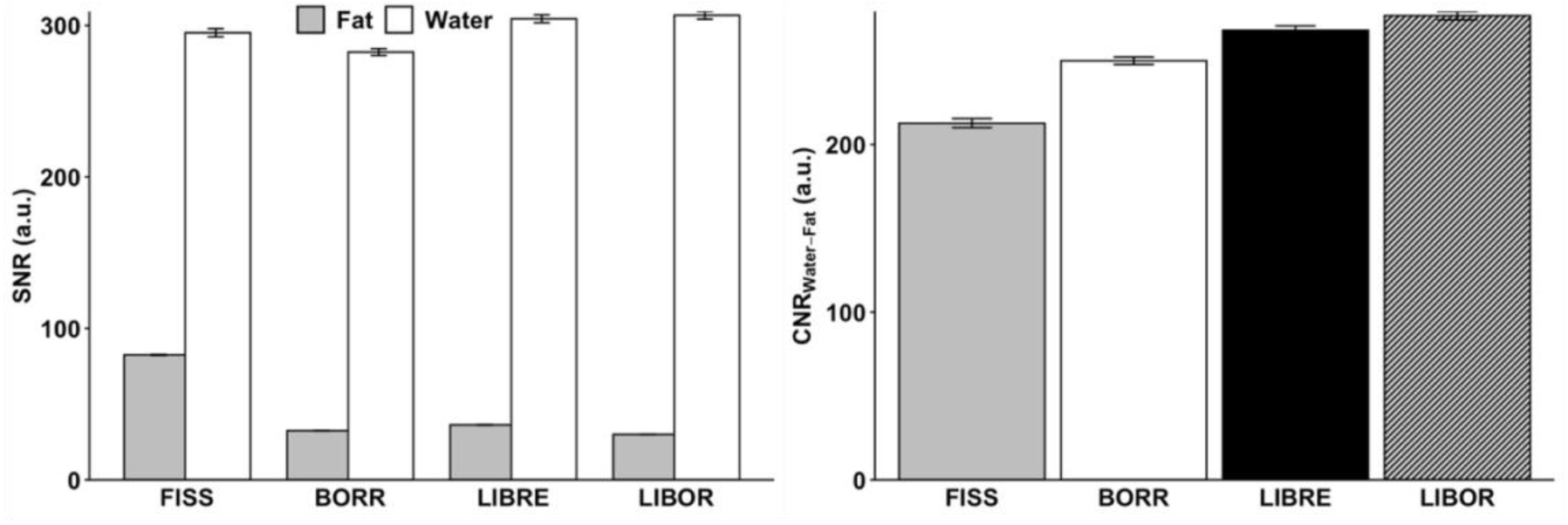
Signal-to-noise ratio (SNR) and contrast-to-noise ratio (CNR) comparison for different fat suppression methods in phantom experiments. (A) Comparison of SNR and (B) CNR between water and fat for four distinct fat signal suppression methods—FISS, BORR, LIBRE, and LIBOR. In the SNR figure, grey denotes fat, and white represents water’s SNR.

In terms of RF energy deposition, SAR values showed that FISS deposited 6.1 times, BORR 3.3 times, and LIBRE 2.5 times more energy than LIBOR (**Figure 4**).

**Figure 4.**
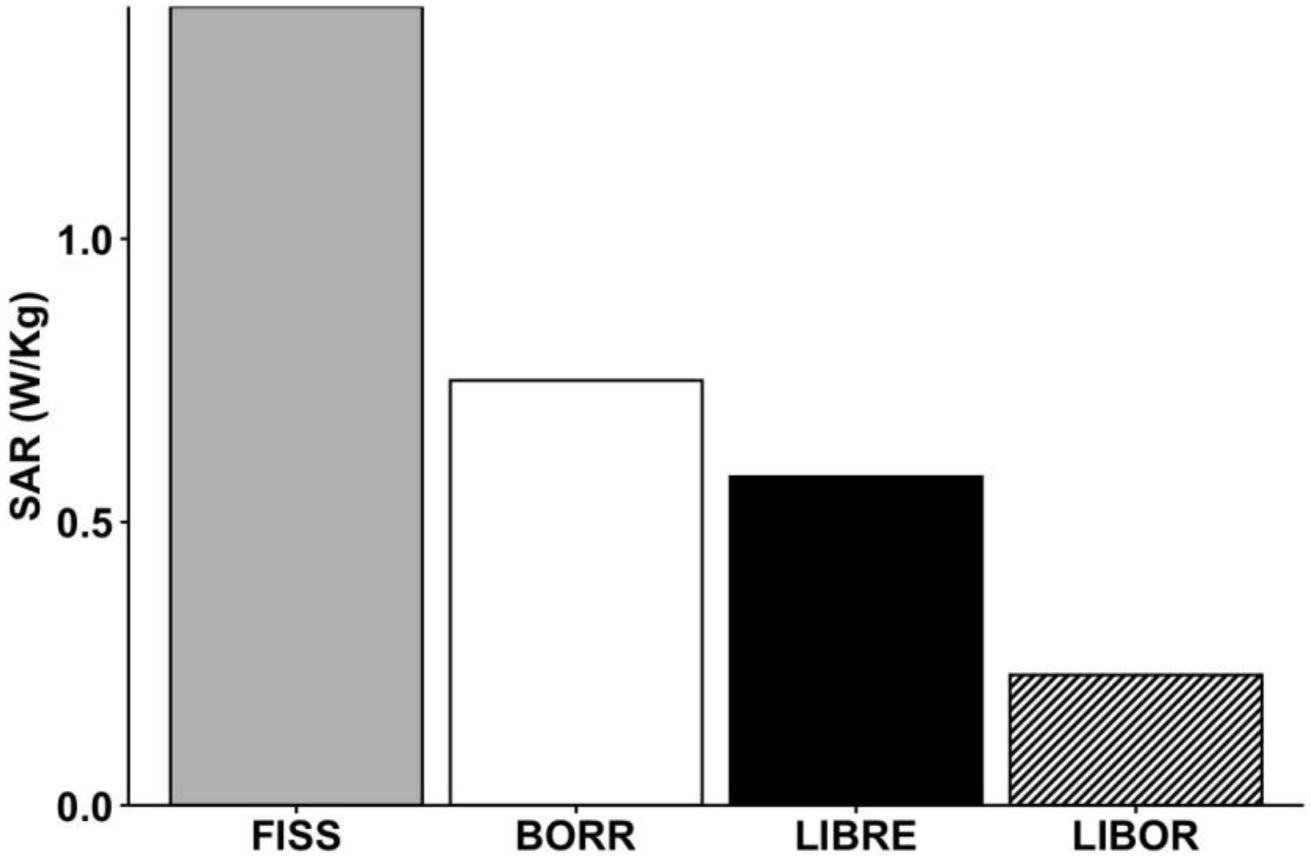
Specific absorption rate (SAR) values for different fat suppression methods in phantom experiments. SAR values for FISS, BORR, LIBRE, and LIBOR in phantom and volunteer measurements. Pulses have identical acquisition parameters summarized in Table 1.

### Fat suppression pulse performance in volunteer experiments using a contrast-free 5D whole-heart free-running bSSFP sequence

In the volunteer study, the performance of each pulse acquisition was visually comparable, with similar brightness levels observed in the myocardium, left ventricular blood pool, chest fat, right lung, and background in the 5D motion-resolved reconstructed images (**Figure 5**). No significant streaking artifacts were observed in the volunteer scans across the sequences with fat suppression, FISS, BORR, LIBRE, and LIBOR.

**Figure 5.**
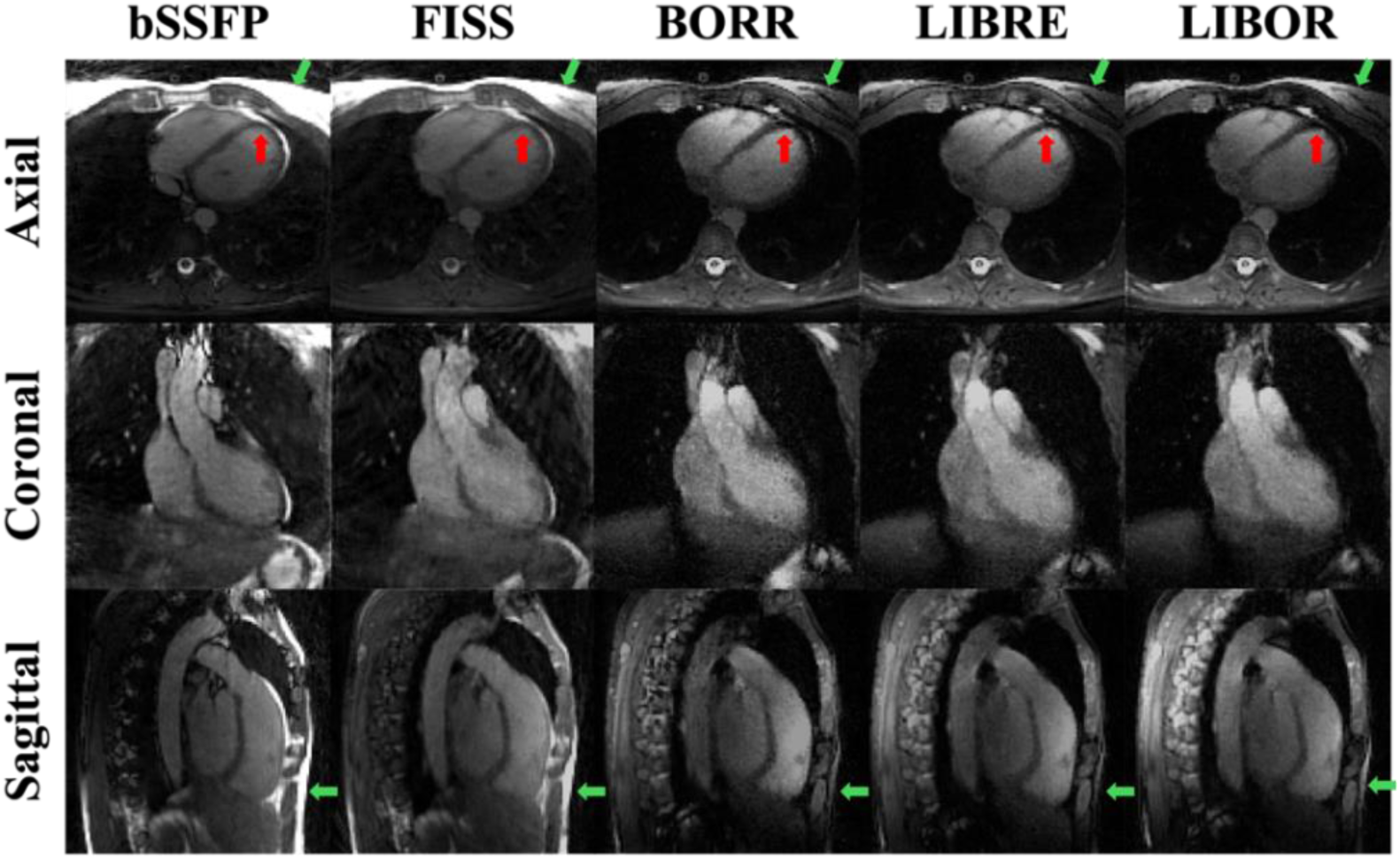
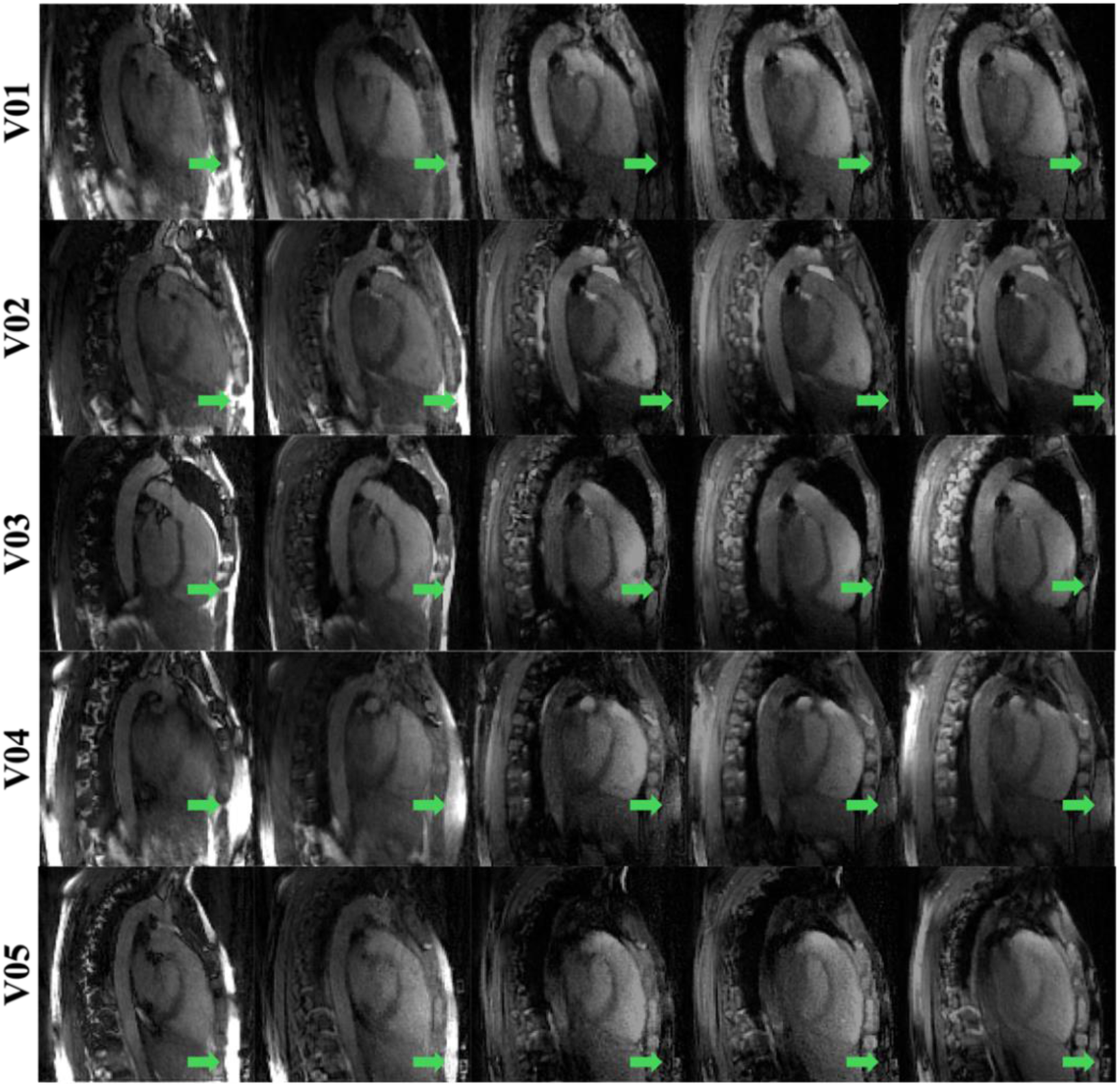
Cardiac and respiratory motion-resolved 5D whole-heart MRI reconstructed for volunteer data acquired with different fat suppression methods. Conventional non-fat-suppressed bSSFP, FISS, BORR-bSSFP, LIBRE-bSSFP, and LIBOR-bSSFP pulses were acquired in 5 volunteers. The images display the heart in the end-diastolic phase of the cardiac cycle and the end-expiration phase of the respiratory cycle. Red arrows show the myocardium wall, and green arrows represent subcutaneous fat. Note that the perceived CNR is slightly reduced compared to previous studies due to the nominal RF excitation angle being limited to 50 degrees in the volunteer experiments. The contrast window and level settings are the same in all images.

Meanwhile, when using the static gridding reconstructed images from the volunteers, BORR exhibited the highest SNR in the ventricular blood pool (17.0 ± 1.5), whereas LIBRE had the lowest value (13.5 ± 1.2). Comparing the SNR for chest fat, BORR had the highest (4.7 ± 2.2), and LIBRE had the lowest (3.6 ± 1.5) values (**Figure 6**). The results showed no significant differences between acquisitions for the SNR of the blood pool (p = 0.565) and myocardium (p = 0.465). However, there was a significant difference in the SNR for chest fat compared to bSSFP (p = 0.0028).

**Figure 6.**
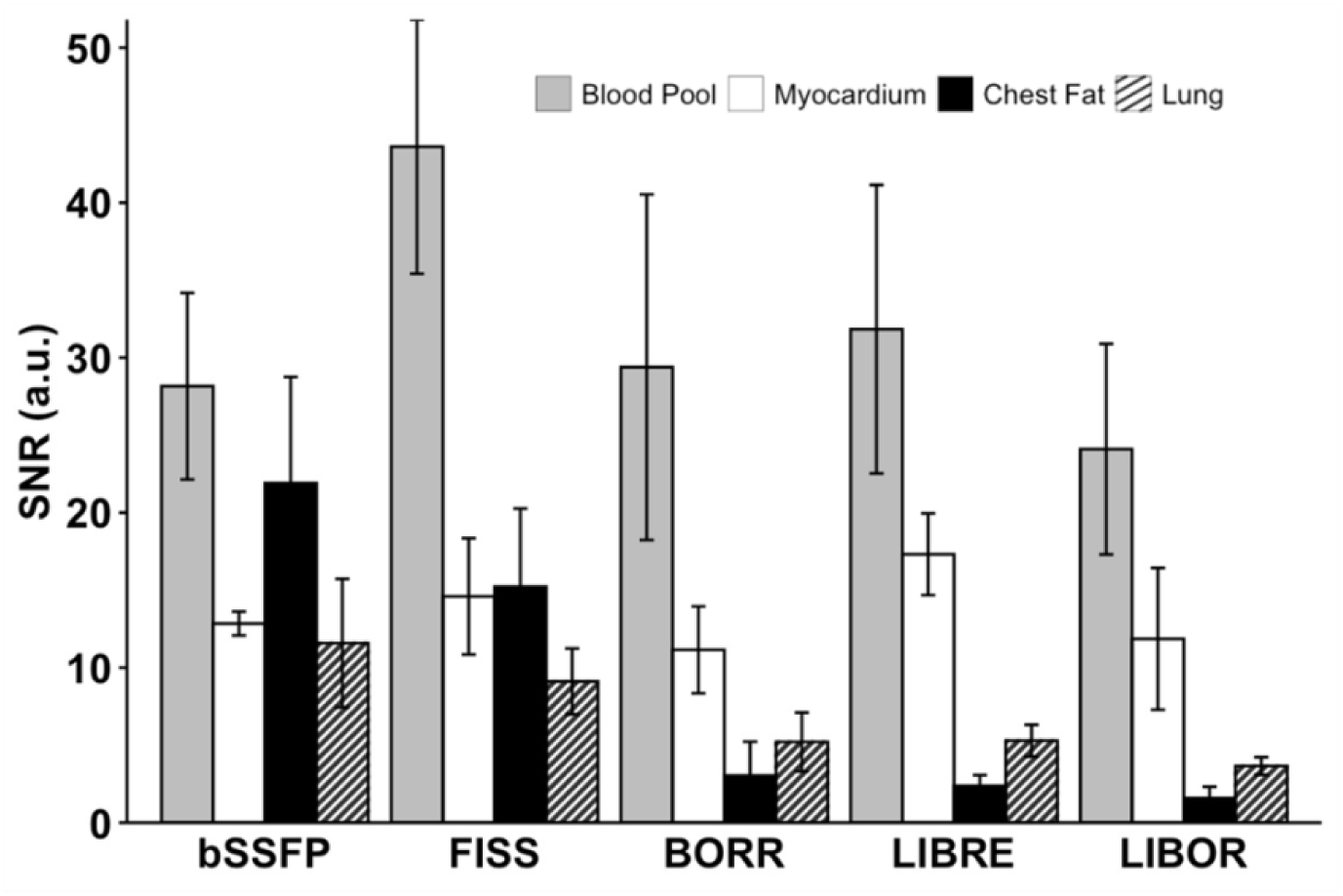
Signal to Noise Ratio (SNR) for volunteer experiments using different pulses. SNR was quantified for blood pool (gray), myocardium (white), chest fat (black), and lung (hatched) using static gridding reconstruction. Regions of interest (ROIs) were manually placed in the septal myocardium, left ventricular blood pool, chest fat, right lung, and background on axial slices. The sequences analyzed include conventional non-fat-suppressed bSSFP, FISS, BORR-bSSFP, LIBRE-bSSFP, and LIBOR-bSSFP pulses. Error bars represent the standard deviation across the cohort.

When comparing the contrast between the blood pool and chest fat, LIBRE displayed the highest CNR_Blood-Fat_ (29.4 ± 9.3), in comparison to bSSFP (6.3 ± 9.1), FISS (28.4 ± 9.6), BORR (26.4 ± 9.3), and LIBOR (22.5 ± 6.8). FISS had the highest CNR_Blood-Myocardium_ (29.0 ± 8.9) compared to bSSFP (15.4 ± 6.1), BORR (18.2 ± 11.4), LIBRE (14.5 ± 9.6), and LIBOR (12.2 ± 8.2) (**Figure 7**).

**Figure 7.**
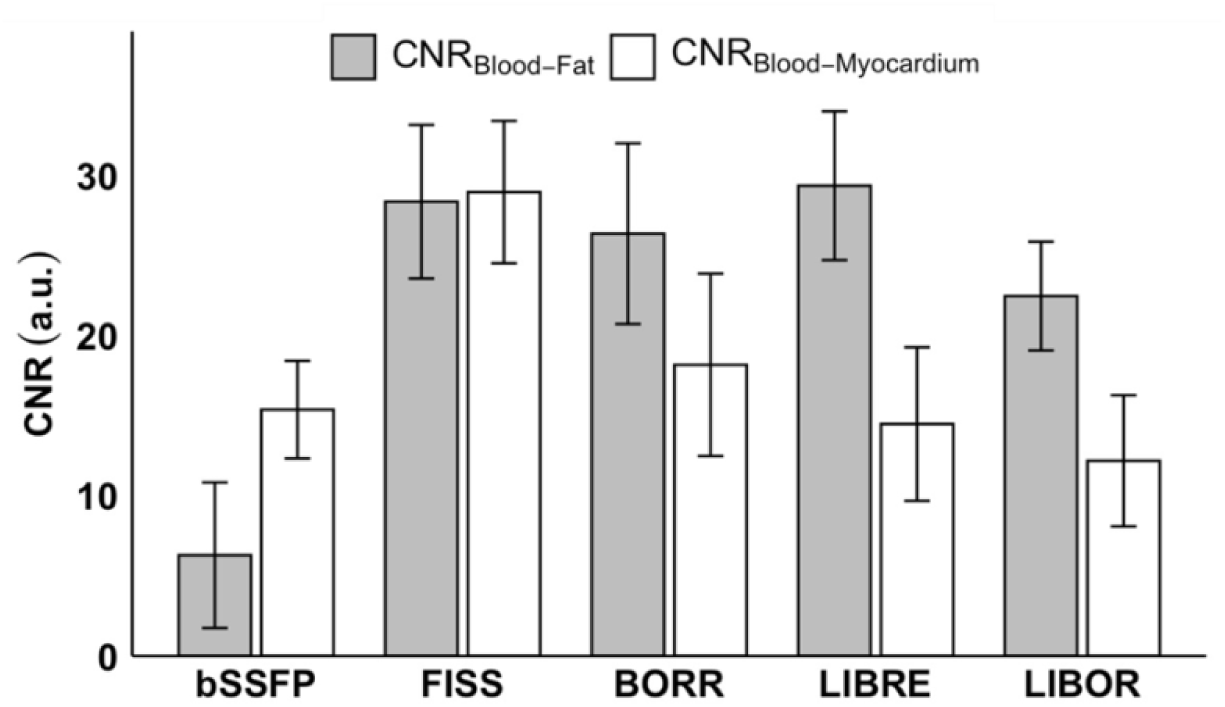
Contrast to Noise Ratios (CNR) for volunteer experiments using different pulse sequences. The CNR_Blood-Fat_ (grey) was calculated by subtracting SNR_Blood Pool_ from SNR_Chest Fat_, and CNR_Blood-Myocardium_ (white) was calculated by subtracting SNR_Blood Pool_ from SNR_Myocardium_. All CNR values were obtained using static gridding reconstruction. The sequences analyzed include conventional non-fat-suppressed bSSFP, FISS, BORR-bSSFP, LIBRE-bSSFP, and LIBOR-bSSFP pulses. Error bars represent the standard deviation across the cohort.

Overall, LIBOR had the lowest SAR (0.26 W.kg^-1^), with 7.0, 7.7, 3.2, and 2.5 times lower than bSSFP (1.81 W.kg^-1^), FISS (1.99 W.kg^-1^), BORR (0.84 W.kg^-1^), and LIBRE (0.65 W.kg^-1^). Additionally, SAR values recorded in volunteers displayed consistent patterns with those observed in phantom experiments **(Figure 8**).

**Figure 8.**
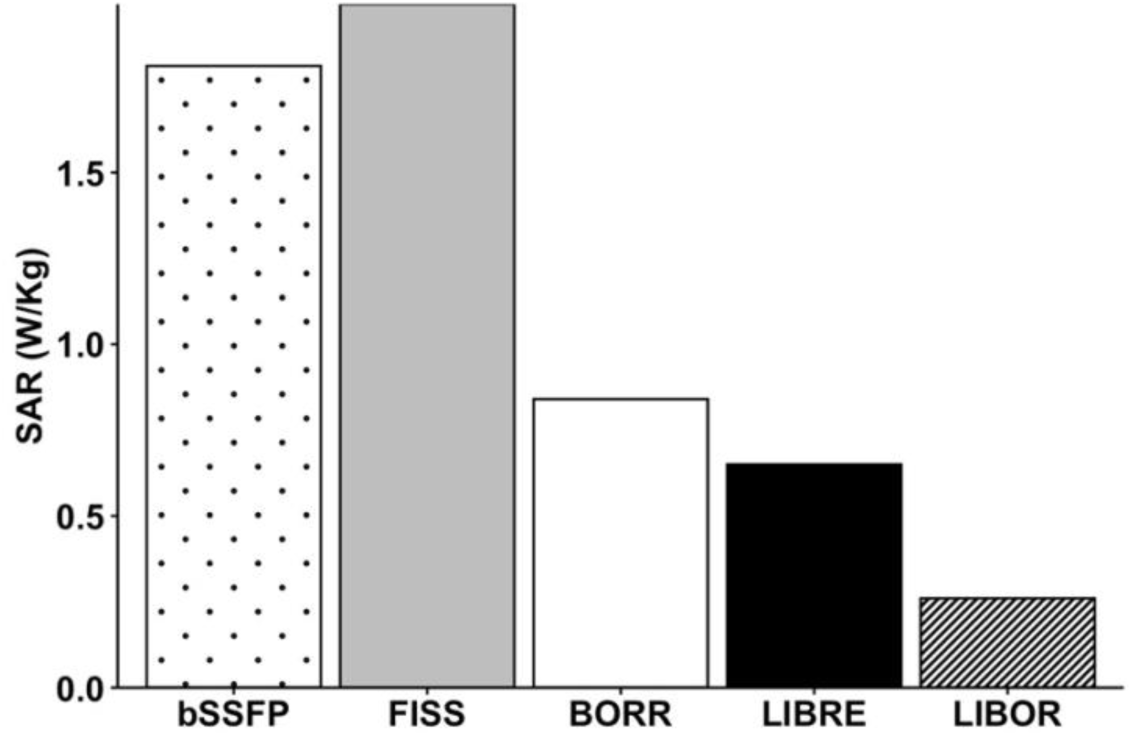
**Specific absorption rate (SAR) values for different fat suppression approaches compared to a bSSFP sequence without any fat suppression in volunteer experiments.**

## DISCUSSION

This study assessed and compared the efficacy of fat signal suppression and energy deposition characteristics (SAR) between off-resonant water excitation RF pulses, such as BORR, LIBRE, and LIBOR, and the repeated interruption of steady-state to suppress fat signal, FISS, in a 3D contrast-free radial whole-heart free-running bSSFP MRI at 1.5T through both phantom and volunteer experiments. Two WE off-resonant RF pulses, BORR and LIBOR, were implemented and tested for the first time at a 1.5T scanner, demonstrating promising results for fat-suppressed 3D contrast-free motion-resolved whole-heart MRI using the free-running framework.

As expected, based on recent findings at 3T [29], the LIBOR pulse resulted in a significant reduction in RF energy deposition both in phantom and volunteer studies while maintaining comparable fat suppression efficacy. This makes using a LIBOR pulse particularly useful for acquisitions requiring a short TR and high SAR intensity, such as free-running sequences based on bSSFP. Its ability to provide robust fat suppression while maintaining low RF power deposition enhances image quality and could potentially improve diagnostic accuracy, particularly in cases where clear differentiation between fat and water signals is crucial, such as fat infiltration or lipomatous hypertrophy, where effective fat suppression can significantly enhance the visibility of subtle pathological features.

Although the BORR pulse resulted in the highest SAR deposit when compared with the off-resonance water excitation pulses, the fat suppression bandwidth of BORR is excellent, which was also observed at 3T [26,29]. The increased SAR deposition of BORR is high because the pulse is inefficient for water excitation, which necessitates an increase in RF power. This, in combination with bSSFP, which is also SAR-intensive, results in reaching the SAR limit. However, using BORR pulses would benefit sequences that are less SAR intensive, for example, sequences that are interrupted by ECG triggering or short sequences, like the typical breath-hold sequences that are performed in CMR. The findings align with previous works on off-resonant pulses like LIBRE and BORR, highlighting their effectiveness in fat suppression and their limitations regarding SAR. This study extends these findings by introducing and validating the LIBOR pulse at 1.5T, addressing the SAR limitations observed with LIBRE and BORR pulses [7,27,29].

The spatial resolution used in this study would be well-suited for cardiac cine imaging, which captures the heart in motion throughout its cardiac cycle. However, due to scan time constraints, incorporating a full stack of 2D cine acquisitions as a reference standard method was not feasible. Cardiac cine imaging is typically used to calculate metrics like ejection fraction, left ventricular volumes, and mass. Previous research has demonstrated the feasibility of these applications within the free-running framework, suggesting potential for future studies to explore this further [41].

Based on visual assessment and comparison with prior work, it was observed that the CNR_Blood-Myocardium_ was not entirely optimal in the volunteer study. This was primarily attributed to having a relatively low nominal RF excitation angle used in this study, which denotes the rotation angle of the water magnetization. Optimal blood-myocardium contrast in bSSFP imaging is typically achieved with RF excitation angles between 60-70 degrees. However, in this study, all nominal RF excitation angles were capped at 50 degrees due to SAR constraints encountered during the BORR acquisition and to ensure a fair comparison among the different methods. This limitation was necessary to avoid exceeding SAR limits, given the pulses used across all sequences. As a result, the observed CNR was a direct consequence of this uniform SAR management strategy.

The lower SAR associated with techniques like LIBRE and LIBOR suggests that they could potentially operate at higher RF excitation angles without exceeding SAR limits, further enhancing the SNR and CNR of the images. Therefore, future studies could explore the performance of these methods under higher nominal RF excitation angles to fully assess their capabilities and achieve higher image quality.

The FISS sequence utilized in this study features short TRs and RF pulses that can lead to a higher SAR due to the concentrated energy distribution. FISS sequence’s design, particularly in a free-running bSSFP framework, effectively balances acquisition speed and resolution while maintaining manageable SAR levels [24]. On the other hand, alternative RF pulses like LIBOR and BORR were evaluated, offering an inherently lower SAR profile. These pulses efficiently distribute RF energy, making them ideal for scenarios requiring extended scan times or higher RF excitation angles without compromising the image quality.

Moreover, a detailed comparison using B0 maps could provide deeper insights into the fat suppression efficacy observed across different sequences, which was beyond the scope of this study. By integrating B0 maps, such as those obtained from multi-echo gradient echo techniques [15], we can accurately assess the true B0 field in each voxel and better characterize the excitation profiles of each pulse. This approach would allow us to understand why fat suppression varied across regions, especially since the FISS sequence has a narrower fat suppression bandwidth compared to WE pulses. Examining how off-resonance effects influenced fat suppression at specific locations could further clarify these observations, providing a more comprehensive understanding of the underlying mechanisms at play.

Confirming prior findings, all RF pulses evaluated in this study show good results, with LIBOR emerging as a promising off-resonant water excitation pulse for fat signal suppression in large FOVs while mitigating SAR limits. It demonstrated potential for functional cardiac imaging at high spatial resolutions similar to FISS and LIBRE [7,24], offering a streamlined workflow compared to conventional CMR protocols within the free-running framework.

By effectively suppressing fat signals, the LIBOR pulse enhances the contrast between the myocardium and surrounding tissues, ensuring clearer visualization of cardiac structures and improving the precision of both qualitative and quantitative evaluations, particularly when high-resolution imaging is required to capture subtle pathological changes or functional abnormalities. Furthermore, in this study, streaking artifacts, which can result from inadequate fat suppression in non-Cartesian readouts, were not prominently observed in the volunteer scans. This outcome reflects the effectiveness of the fat suppression methods, including FISS, BORR, LIBRE, and LIBOR, in managing fat signal interference. However, subtle differences in artifact severity between sequences were assessed qualitatively through visual inspection of the reconstructed images. Future studies could incorporate more quantitative measures, such as artifact power or structured noise metrics, to objectively evaluate and compare the performance of different sequences in mitigating artifacts.

### Limitations

In this study, several limitations should be considered when interpreting the findings. Firstly, the spatial resolution in this study was limited by the need to balance total scan duration and the restricted TR range of 2.2 to 3.0 ms required for the FISS sequence at 1.5T [24]. This TR constraint limited the achievable resolution to around 2 mm³ for practical acquisition times. However, this spatial resolution is insufficient for clearly visualizing coronary arteries, a key application for the bSSFP free-running sequence. Despite this limitation, the primary goal of comparing fat suppression techniques was met effectively. Previous research, including earlier work on coronary MRA, has demonstrated that higher resolutions are possible and effective for coronary imaging using bSSFP free-running sequences [6,7,24]. Although not explored in this study, future research with higher spatial resolution could benefit coronary artery imaging.

As detailed in the Methods section, the motion-compensated reconstruction used compressed sensing techniques with regularization in the cardiac and respiratory dimensions, following the framework described by Di Sopra *et al.* [39]. This approach effectively suppressed noise and artifacts while maintaining spatial and temporal resolution, enabling the generation of 5D motion-resolved images for qualitative assessment. However, the reconstruction process was time-consuming, indicating a need for a faster and more efficient reconstruction code. Also, having a larger pool of volunteers and validation on patient cohorts would be of interest to further validate these results. Additionally, this study was conducted on a cohort of healthy, young volunteers with low BMI (mean weight 66.4 kg). While this controlled setup allowed for systematic comparisons, it may not fully capture the challenges posed by higher fat content in patients with obesity or heart disease. However, the suppression bandwidth of off-resonance pulses is relatively large, and no issues are foreseen, as has been demonstrated in patient cohorts utilizing LIBRE pulses at 3T, where field inhomogeneities are larger [34–36]. Nevertheless, future studies should include a broader patient cohort to assess the robustness of these methods in clinical settings with varying body compositions.

## CONCLUSION

Different off-resonance water-excitation methods were implemented and compared in fat-suppressed contrast-free 5D whole-heart free-running bSSFP cardiac MRI at 1.5T, marking the first report on both LIBOR and BORR pulses. While maintaining a similar and adequate fat suppression and having a short TR with sufficient blood-muscle contrast, LIBOR deposited the minimum RF power compared to other pulses, ensuring optimal image quality for future clinical applications.

## Abbreviations

2D: Two-Dimensional
ADMM: Alternating Direction Method of Multipliers
BORR: Binomial Off-Resonant Rectangular
bSSFP: balanced Steady-State Free Precession
CMR: Cardiac Magnetic Resonance
CNR: Contrast-to-Noise Ratio
CS: Compressed Sensing
DICOM: Digital Imaging and Communications in Medicine
FISS: Fast Interrupted Steady-State
FOV: Field of View
GRE: Gradient Recalled Echo
LIBOR: Lipid Insensitive Binomial Off-Resonant
LIBRE: Lipid Insensitive Binomial Off-Resonant RF Excitation
LLR: Local Low-Rank
RF: Radiofrequency
ROI: Region of Interest
SAR: Specific Absorption Rate
SNR: Signal-to-Noise Ratio
TE: Echo Time
TR: Repetition Time
TV: Total Variation
WE: Water Excitation

## Data Availability

All data produced are available online at Zenodo with DOI number of: 10.5281/zenodo.13868461

https://doi.org/10.5281/zenodo.13868461

## Acknowledgments

This study was supported by funding from the Swiss National Science Foundation (SNSF), including grants PCEFP2_194296 attributed to JB and 320030_173129 and 320030B_201292, which provided essential support for the development of the free-running framework, including the reconstruction used in this study. The development of the reconstruction framework was also partly funded by the SNSF grant 310030_215604, for which JY is the principal investigator. CG received funding from the SNSF, InnoSuisse, Center for Artificial Intelligence in Medicine (CAIM) at the University of Bern, the GAMBIT Foundation, the Novartis Foundation for Medical-Biological Research, and the Swiss Heart Foundation, all outside the scope of the submitted work. Additionally, CG serves as Editor-in-Chief of *The International Journal of Cardiovascular Imaging*, Springer.

